# Left Ventricular Diastolic Intraventricular Pressure Differences and Propagation Velocity with 4D-Flow MRI in Patients with Hypertrophic Cardiomyopathy

**DOI:** 10.64898/2026.10.15.26365692

**Authors:** Luz V. Stipechi, Damian Craiem, Umit Gencer, Mariano E. Casciaro, Jean-Sébastien Hulot, Albert Hagege, Gilles Soulat, Elie Mousseaux

## Abstract

**Background:** Transthoracic echocardiography (TTE) remains the gold standard for assessing left ventricular (LV) diastolic dysfunction (DD), but noninvasive alternatives are needed to overcome geometric and acoustic limitations. 4D-Flow MRI enables comprehensive assessment of intraventricular pressure differences (IVPDs) and flow propagation velocity (VP). We aimed to evaluate 4D-Flow-derived IVPDs and VP in patients with hypertrophic cardiomyopathy (HCM).

**Methods:** Thirty-eight HCM patients and 38 age- and sex-matched healthy controls underwent TTE and 4D-Flow MRI. Spatiotemporal maps of velocity and IVPDs, decomposed into inertial and convective components, were derived along the main inflow streamline from the mitral valve to the apex. Peak early filling rate (PEFR) was used to normalize IVPDs and VP.

**Results:** While absolute peak flow rates were comparable, HCM patients exhibited significantly faster VP (100.4 vs. 60.2 cm/s, p < 0.01) and higher positive early E IVPD (1.40 vs. 0.92 mmHg, p < 0.01) compared to controls. Normalizing IVPDs by PEFR amplified group differences, revealing a markedly increased flow-normalized E-IVPD (5.13 vs. 2.94 × 10⁻³ mmHg·ml⁻¹·s, p < 0.001) and deeper reverse ER IVPD (-6.06 vs. -4.61 × 10⁻³ mmHg·ml⁻¹·s, p < 0.01) in HCM. These differences were primarily driven by the inertial components. Multivariable analysis demonstrated that ER IVPD is independently determined by PEFR and the TTE E/E’ ratio (r = 0.65, p < 0.001).

**Conclusions:** 4D-Flow MRI-derived, flow-normalized IVPDs and propagation velocity provide robust, noninvasive markers of altered LV chamber stiffness and impaired diastolic filling in HCM, offering deeper 3D hemodynamic insights into diastolic dysfunction.

**Visual Abstract:** 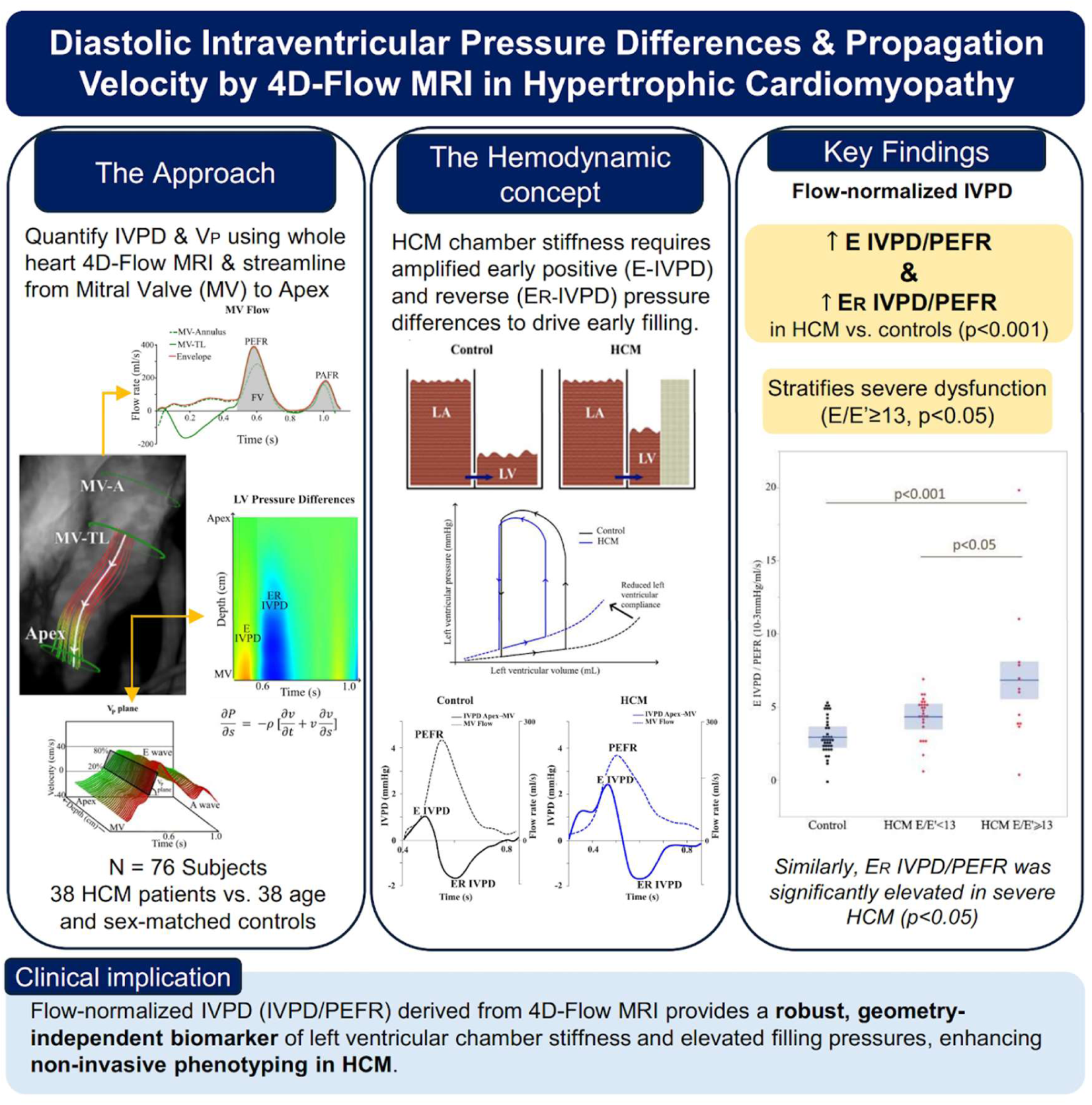

## Introduction

Heart failure, a major driver of cardiovascular morbidity and mortality, fundamentally results in reduced cardiac output and/or elevated left ventricular (LV) filling pressures at rest or during exercise. Accurate assessment of these filling pressures and their pathophysiological determinants is essential for phenotyping patients with heart failure, diastolic dysfunction (DD), and cardiomyopathies at risk of progressing to overt heart failure. To date, transthoracic echocardiography (TTE) remains the clinical gold standard for assessing DD. Current diagnostic algorithms rely on combining parameters such as early (E) and late (A) peak mitral filling velocities, early peak longitudinal velocity of the mitral annulus (E’), and left atrial (LA) size to estimate LV filling pressures[1]. However, the weak correlation often observed between Doppler echocardiographic estimates and invasive catheterization measurements reinforces the need for more robust noninvasive parameters[2,3]. Fluid dynamics theory, applied to color Doppler M-mode (CDMM) echocardiography, has been proposed as a method to assess intraventricular pressure difference (IVPD) and flow propagation velocity (V_P_) to better characterize LV filling pressures[4,5].

While the simplified Bernoulli equation is effective for estimating spatial pressure difference across restrictive orifices in stenotic or regurgitant lesions, it is inaccurate for unrestrictive flows where inertial forces are not negligible[6,7]. Based on Euler equation[5,8], the spatial pressure difference is the net result of the change in velocity over time at a given spatial point (inertial component, ∂v/∂t) and the change in velocity with respect to spatial position at any instant (convective component, v·∂v/∂s). Despite their theoretical advantages, Euler equation-based methods remain underutilized in routine echocardiography due to limited technical feasibility and the complex pathophysiological interpretation of the results[6,7]. Furthermore, CDMM is constrained by acoustic window dependency and accurately captures velocities only parallel to the ultrasound beam[6]. These issues can introduce significant errors in the estimation of IVPD and V_P_ due to insufficient sampling of the blood velocity field. Consequently, when spatial or temporal derivatives are applied to under-sampled data, noise amplification is inevitable.

Two-dimensional (2D) phase-contrast magnetic resonance imaging (PC-MRI) is an alternative technique for evaluating velocities and flow rates over time and space along or across streamlines. In conditions of highly variable flow, relative pressure differences within a single cavity or across normal and stenotic valves have been validated using 2D PC-MRI, both computationally and experimentally[6,9,10]. However, similar to CDMM, velocity encoding in 2D PC-MRI remains intrinsically dependent on plane orientation. Given the asymmetric and complex nature of LV inflow, evaluating DD using time-resolved, three-dimensional velocity data is highly preferable[9,11,12]. By capturing velocities across the entire LV inflow tract, 4D-Flow MRI yields peak velocity measurements that are highly concordant with those from TTE[12,13]. Furthermore, LV filling parameters based on peak flow, as opposed to peak velocities across the mitral valve, demonstrate stronger associations with aging[12] and prognosis in patients with hypertrophic cardiomyopathy (HCM), who often present with diastolic dysfunction[14]. Finally, the ability to derive V_P_ using 4D-Flow MRI has been recently demonstrated [12].

The present study aims to demonstrate the feasibility of a 4D-Flow MRI-based method for characterizing early diastolic IVPDs and V_P_. We compared these 4D-Flow markers against established echocardiographic indices of elevated LV diastolic pressures in a population with HCM vs controls.

## Materials and methods

### Study Cohort

This study was part of the French research project “Registry of hypertrophic cardiomyopathy” (REMY) (*ClinicalTrials.gov* identifier: NCT01091480)[15]. Launched at 26 centers in 2016, REMY is a large-scale, ongoing, nationwide, prospective, multicenter observational study focusing on the diagnosis, management, and prognosis of sarcomeric and non-sarcomeric HCM. This registry included prospectively adult participants (aged ≥ 16 years) with HCM, defined as LV hypertrophy (maximum wall thickness ≥15 mm in sporadic cases, or ≥13 mm with a family history of HCM or a positive genetic test) unexplained by abnormal loading conditions. However, the study presented here is an exploratory substudy conducted by a single investigator (E.M.) from one of these centers, who recruited 44 participants between June 2018 and June 2022 to undergo a cardiac MRI including a 4D-Flow sequence. Six patients had to be excluded either because the 4D flow did not fully cover the left ventricle or because the scans were performed on a 3-Tesla system. The remaining 38 HCM patients were matched by age and sex to 38 healthy volunteers with normal electrocardiograms and no known cardiovascular disease, serving as a control group. Following informed consent, all participants underwent TTE and cardiac MRI within a 48-hour window. Echocardiographic examinations were performed according to current societal guidelines[1].

### MRI examinations

All cardiac MRI examinations were performed with a 1.5-T scanner (SIGNA Artist, GE healthcare). LV volume, mass, and ejection fraction were measured from 2D steady-state free precession LV short-axis cine stacks[16]. Inversion recovery late gadolinium enhancement (LGE) images were obtained 12 minutes after intravenous injection of 0.2 mmol/kg gadopentetate dimeglumine (Dotarem; Guerbet). The LGE extent was quantified using a threshold of ≥6 standard deviations above the mean signal of remote normal myocardium and expressed as a percentage of LV mass. Left atrial volumes (LAV) were assessed using the biplane area–length method from 2- and 4-chamber cine views at LV end-diastole and end-systole. All cavity volumes were indexed to body surface area (BSA). Derived structural indices were calculated including the LV remodeling index defined as the ratio of LV mass to LV end-diastole volume (LVM/LVEDV) and the Left Atrioventricular Coupling Index (LACI), defined as the ratio of LAV at end-diastole to LVEDV[17]. All these parameters were analyzed using Cvi42 (Circle CVI). Whole-heart 4D-Flow MRI was acquired using a free-breathing, retrospective sequence without a respiratory navigator for six to nine minutes, immediately following contrast injection. Scanning parameters included: velocity-encoding value of 160 cm × sec^−1^ in all directions, acceleration factor of 4.0 (Kt-ARC), mean acquired spatial resolution of 2.2 × 2.2 × 2.2 mm, flip angle of 15°, repetition/echo time of 4.2/1.7, bandwidth of 62 kHz, two views per segment, and a temporal resolution of 33.6 msec reconstructed into 50 frames per cardiac cycle.

### Image processing

Post-processing of 4D-Flow images was performed using in-house software (www.lattido.com) developed in C# by Universidad Favaloro (Buenos Aires, Argentina) in collaboration with Université de Paris[18]. Eddy current-induced background phase offsets were automatically corrected using a third-order surface interpolation[19]. Measurements began with the manual drawing of two parallel regions of interest (ROIs) within the LV volume (Figure 1A): one at the level of the mitral valve tip leaflets (MV-TL) and the second located 4 cm distally towards the apex. These ROIs were positioned orthogonally to the main incoming flow direction at peak E-wave by a trained operator (U.G.), using a streamline tool which dynamically visualized blood velocity. ROIs were manually adjusted during key diastolic phases, while intermediate temporal phases were automatically calculated via temporal interpolation to seamlessly cover the entire cardiac cycle[18]. To account for longitudinal atrioventricular plane displacement, a third ROI was drawn at the mitral valve annulus (MV-A).

**Figure 1:**
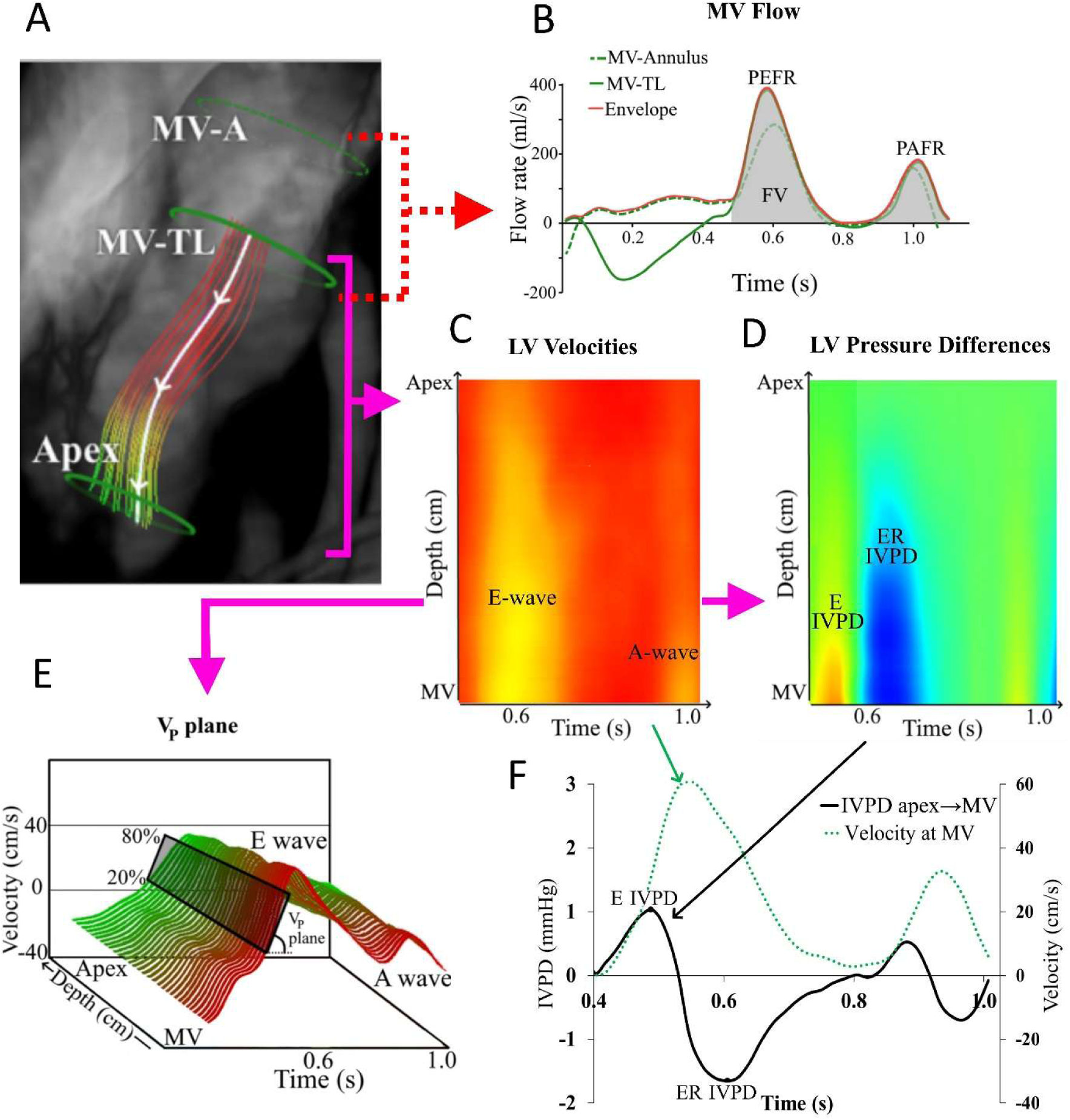
Illustration of the method used to quantify transmitral flow, propagation velocity (V_P_), and intraventricular pressures differences (IVPDs) based on 4D-Flow data. Figure A shows the three ROIs at the mitral valve annulus (MV-A), at the tips of the mitral valve leaflets (MV-TL), and the left ventricular apex, along with the main inflow streamline. Figure B displays the flow rates obtained at the MV-A and MV-TL levels. The red curve represents the merged maximum flow envelope between these two levels, used to estimate the peak early filling rate (PEFR), peak atrial filling rate (PAFR) and filling volume (FV). Figures C and D show the velocity and pressure difference maps between the MV-TL (bottom) and the apex (top) over time. Figure E illustrates the method for calculating V_P_ wherein a 3D time-space plane is fitted to the upslope portion of the velocity curves between the MV-TL and the apex. Figure F displays the superimposed temporal velocity and IVPD curves derived from the maps. E IVPD and ER IVPD denote the peak positive and reversal pressure differences, respectively.

### Definition of flow and velocity filling parameters

As shown in Figure 1B, the LV flow filling parameters were derived by calculating the envelope of a merged flow rate curve (red line), combining data from the MV-A (dotted line) and MV-TL (solid line) ROIs. LV filling volume (FV) was calculated as the area under this merged curve as explained elsewhere[14]. Peak early filling rates and peak atrial filling rates (PEFR and PAFR) were identified as the maximum flow values across the MV during early LV filling and atrial contraction, respectively. PEFR was normalized to FV (PEFR/FV), providing a standardized indicator of LV filling to allow inter-subject comparison regardless of baseline stroke volume[13]. The PEFR/PAFR ratio was calculated using peak flow measurements. Velocity vectors with the maximum magnitude within the MV-TL ROI were measured at the peak of both the E-wave (E_TL_) and A-wave (A_TL_), to further calculate the E/A_TL_ velocity ratio.

### Time-space velocity color map

Analogous to the CDMM echocardiography[20], the spatiotemporal distribution of blood flow velocities inside the LV was mapped from the MV to the apex across 50 time phases as explained elsewhere[12]. Figure 1C displays the time-space velocity color map: (x-axis: time; y-axis: distance from MV to apex). The complex LV inflow pattern was simplified into a single curvilinear pathway representing the core velocity trajectory during diastolic filling. This was achieved by propagating streamlines from the MV-ROI at peak E-wave until 20% of them reached the apical ROI. These streamlines were spatially averaged, and the resulting smooth curve (Figure 1A, white curve) served as the primary velocity trajectory. Velocity magnitudes were sampled at 0.5 mm intervals along this streamline to generate the velocity spatiotemporal color map.

### Estimation of intraventricular pressure difference (IVPD)

Three-dimensional blood flow through the LV is governed by the Navier-Stokes equations for incompressible fluids,

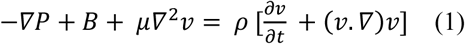

where, *v* represents velocity, *P* is pressure, *B* is the body forces acting on the fluid (such as gravity), *ρ* is blood density and *μ* is viscosity. Assuming viscous friction is negligible and gravitational forces are balanced by hydrostatic buoyancy (*B*=0), restricting the analysis to the main streamline reduces the governing principles of Equation 1 to the 1D Euler equation:

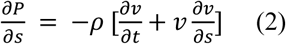

where *s* represents the spatial distance along the pathway from the MV to the apex. Consequently, the pressure gradient represents the net sum of local inertial acceleration (*∂v/∂t*) and convective acceleration (*v.∂v/∂s*), multiplied by blood density (*ρ*=1060 kg/m^3^). Intra-ventricular pressure gradients were numerically estimated to create a time-space pressure gradient color map (Figure 1D). The total IVPD between the MV-TL and the apex was calculated via direct spatial integration along the streamline:

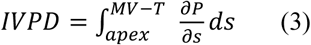

To estimate the pressure difference along the streamline using Eq. 3, the integration path was divided into *k* segments of length Δs. The total pressure drop between the apex and the MV-TL was then calculated using the direct integration method:

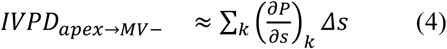

Figure 1F illustrates a representative total IVPD curve over time between LV apex and MV-TL. During the E-wave, the maximum positive (E IVPD) and minimum reversal (ER IVPD) values were extracted. Since the inertial and convective components of the IVPD can also be decomposed separately using Eq. 2, three additional IVPD values were measured: the maximum positive inertial IVPD (E_inertial_IVPD), the minimum reversal inertial IVPD (ER_inertial_IVPD) and the minimum reversal convective IVPD (E_convective_IVPD).

### Velocity propagation (V_P_)

The flow propagation velocity (V_P_) was estimated using an approach adapted from thoracic aortic pulse wave velocity assessments, previously detailed in a previous study[12]. Briefly, a 3D time-space plane was fitted to the upslope portion (20–80%) of the velocity curves between the MV and apex (Figure 1E). The slope of this fitted plane provided a direct measure of V_P_. To adjust for varying LV filling magnitudes, both IVPDs and V_P_ were normalized by PEFR.

### Statistical Analysis

Continuous and categorical data are expressed as mean±SD, median [interquartile range] or frequencies (percentages). Subsequently, based on recommendation for the evaluation of LV diastolic function[1], the HCM cohort was stratified based on echocardiographic E/E’_TTE_ ratio: Subgroup 1 (E/E’<13) and Subgroup 2 (E/E’≥13). Differences between the control group and the HCM group, as well as between the two HCM subgroups, were assessed using a Student’s t-test, a Welch’s t-test, a Mann-Whitney U-test, a Kruskal-Wallis test, or a Fisher’s exact test, as appropriate. Differences between controls and the two HCM subgroups were further assessed using one-way ANOVA with post hoc Wilcoxon testing. Univariate Pearson correlations were used to explore the determinants of positive and reversal IVPDs. Multivariable models were then constructed using forward stepwise variable selection, incorporating all significant univariate predictors alongside age. Additionally, the HCM cohort was dichotomized based on median LAVi (50 ml/m²), LACI (0.4) and BNP (186 ng/L) to compare IVPD measurements. Interobserver agreement (U.G. and L.V.S.) for IVPD and V_P_ measurements was assessed in a random subset (10 controls, 10 HCM) using the intraclass correlation coefficient (ICC) and the coefficient of variation (CoV). A p-value < 0.05 was considered statistically significant. All analyses were performed using JMP 10.0 (SAS Institute).

## Results

Table 1 outlines the baseline demographic and clinical characteristics of the study cohort. Compared to age- and sex-matched controls, patients with HCM had significantly higher body weight, body mass index, and BSA (all p<0.01). Within the HCM cohort, 15 patients (39%) had hypertension (p<0.001), 23 (61%) were classified as NYHA class ≥ 2 (p<0.001), 9 (24%) had a history of atrial fibrillation (p<0.001), and the median BNP level was 186 [83–415] ng/L.

**Table 1:** Demographic and baseline characteristics in control group vs HCM group and between subgroups of HCM based on an TTE E/E’ cutoff of 13.

|  | Control Group | HCM group | p | HCM group 1<br>E/E'<13 | HCM group 2<br>E/E'≥13 | p |
| --- | --- | --- | --- | --- | --- | --- |
| Number | 38 | 38 |  | 24 | 14 |  |
| **Clinical characteristics** |  |  |  |  |  |  |
| Age, Years | 55 ± 18 | 52 ± 19 | 0.529 | 50 ± 17 | 56 ± 22 | 0.310 |
| Female, n (%) | 12 (32) | 12 (32) | 1.00 | 6 (25) | 6 (43) | 0.257 |
| Weight, kg | 67.1 ± 10.3 | 77.7 ± 17.9 | <0.01 | 83.6±17.3 | 67.5±14.3 | <0.01 |
| Height, cm | 169.0 ± 8.8 | 170.9 ± 8.3 | 0.314 | 172.6±8.1 | 168.0±7.1 | 0.088 |
| Body mass index, kg/m<sup>2</sup> | 23.5 ± 2.7 | 26.6 ± 6.0 | <0.01 | 28.1±6.1 | 23.9±4.9 | <0.05 |
| Body surface area, m<sup>2</sup> | 1.77 ± 0.17 | 1.91 ± 0.25 | <0.01 | 1.99±0.23 | 1.77±0.21 | <0.01 |
| Hypertension, n (%) | 0 (0) | 15 (39) | <0.001 | 9 (38) | 6 (43) | 0.745 |
| History of atrial fibrillation, n (%) | 0 (0) | 9 (24) | <0.001 | 6(25) | 3(21) | 0.802 |
| NYHA functional class ≥ 2, n (%) | 0 (0) | 23 (61) | <0.001 | 12(50) | 11(79) | 0.802 |
| BNP, ng/L | - | 186 [83-415] | - | 126 [56-198] | 415 [225-1045] | <0.01 |
| Maron classification Type I/II/III/IV, n(%) | - | 7(17)/15(41)/3(34)/3(8) | - |  |  |  |

HCM: Hypertrophic cardiomyopathy, TTE: Transthoracic echocardiogram, NYHA: New York heart association functional classification, BNP: B-type natriuretic peptide. Continuous data are expressed as mean±SD or median [interquartile] values.

Table 2 summarizes conventional TTE and Cardiac MRI parameters, along with derived indices. Echocardiographic early filling velocity (E_TTE_) was slightly higher in the HCM group than in the control group (p<0.05), whereas the tissue Doppler early diastolic velocity (E’_TTE_) was lower (p<0.001) driving a substantially elevated E/E’_TTE_ ratio in HCM patients (11.4 vs 6.4, p<0.001). The E/A_TTE_ ratio remained comparable between groups. Ten HCM patients (26%) exhibited a left ventricular outflow tract (LVOT) gradient >30 mmHg (median gradient 53 [40-105] mmHg). Tricuspid regurgitation (TR) velocity ≥280 cm/sec was also present in 7 (18%) HCM patients. While MRI-derived LV volume indices and ejection fractions were equivalent between groups, severe structural remodeling was evident in the HCM group, highlighted by elevated LVMi, LVM/EDV ratio, LAVi, and LACI (all p<0.001). LGE was present in 28 (74%) of HCM patients, though only 7 (18%) had an LGE extent >15% of LV mass.

**Table 2:** Basic TTE and MRI parameters with derived indices in control group vs HCM group and between subgroups of HCM based on an TTE E/E’ cutoff of 13.

|  | Control Group | HCM group | p | HCM group 1<br>E/E'<13 | HCM group 2<br>E/E'≥13 | p |
| --- | --- | --- | --- | --- | --- | --- |
| Number | 38 | 38 |  | 24 | 14 |  |
| **TTE measurements** |  |  |  |  |  |  |
| E<sub>TTE</sub>, cm/s | 66.5 ± 16.2 | 79.1 ± 24.6 | <0.05 | 71.6±22.4 | 91.8±22.8 | <0.05 |
| A<sub>TTE</sub>, cm/s | 59.8 ± 22.2 | 63.7 ± 30.7 | 0.545 | 57.3±18.8 | 75.6±43.7 | 0.189 |
| E/A<sub>TTE</sub> | 1.24 ± 0.50 | 1.45 ± 0.81 | 0.207 | 1.31±0.56 | 1.71±1.12 | 0.271 |
| E'<sub>TTE</sub>, cm/s | 12.5 ± 5.4 | 8.1 ± 3.8 | <0.001 | 9.6±3.9 | 5.5±1.6 | <0.001 |
| E/E'<sub>TTE</sub> | 6.4 ± 2.9 | 11.4 ± 5.5 | <0.001 | 8.0±2.7 | 17.1±4.2 | <0.001 |
| TR velocity ≥280 cm/s, n (%) | 0 (0) | 7 (18) | <0.001 | 3 (13) | 4 (29) | 0.225 |
| Left atrium diameter, mm | 33.5 ± 5.5 | 42.4 ± 6.4 | <0.001 | 42.5±5.9 | 42.3±7.3 | 0.940 |
| LVOT obstruction (>30mmHg), n (%) | 0 | 10 (26) | <0.001 | 5(21) | 5(36) | 0.184 |
| LVOT gradient in all HCM, mmHg | - | 0 [0-40] | - | 0[0-28] | 0[0-66] | 0.231 |
| LVOTO gradient (n=10) mmHg | - | 53 [40-105] | - | 51[45-98] | 55[38-110] | 0.231 |
| **MRI measurements** |  |  |  |  |  |  |
| LVEDVi, ml/ m<sup>2</sup> | 73.3 ± 15.9 | 75.5 ± 20.8 | 0.594 | 72.8±16.3 | 80.2±27.0 | 0.361 |
| LVESVi, ml/ m<sup>2</sup> | 27.8 ± 7.5 | 31.3 ± 17.3 | 0.261 | 28.9±13.0 | 35.3±23.0 | 0.355 |
| Stroke volume, ml | 80.6 ± 19.8 | 84.7 ± 25.1 | 0.435 | 87.7±24.8 | 79.7±25.7 | 0.356 |
| LVEF, % | 61.7 ± 5.0 | 62.3 ± 12.8 | 0.784 | 64.2±10.5 | 58.9±16.0 | 0.283 |
| LVMi, g/ m<sup>2</sup> | 56.0 ± 9.4 | 100.7 ± 33.5 | <0.001 | 91.8±27.8 | 115.9±37.8 | <0.05 |
| LVM/LVEDV, g/ml | 0.79 ± 0.19 | 1.39 ± 0.49 | <0.001 | 1.29±0.42 | 1.54±0.57 | 0.169 |
| LAVi, ml/m<sup>2</sup> | 36.1 ± 11.7 | 55.4 ± 30.3 | <0.001 | 47.4±24.9 | 69.2±34.5 | 0.050 |
| LACI | 0.25 ± 0.11 | 0.53 ± 0.43 | <0.001 | 0.43±0.36 | 0.70±0.49 | 0.091 |
| Presence of LGE, n (%) | 0 (0) | 28 (74) | <0.001 | 17 (71) | 11 (79) | 0.598 |
| LGE extent, % of LVM | 0 (0) | 8.3 ± 8.3 | <0.001 | 6.9±7.1 | 10.6±11.1 | 0.276 |
| LGE extent ≥ 15%, n (%) | 0 (0) | 7 (18) | <0.001 | 3 (13) | 4 (29) | 0.225 |

HCM: Hypertrophic cardiomyopathy, TTE: Transthoracic echocardiogram, TR: Tricuspid regurgitation, LVOTO: Left ventricular outflow tract obstruction, LVEDVi: Left ventricular end-diastolic volume indexed, LVESVi: Left ventricular end-systolic volume indexed, LVEF: Left ventricular ejection fraction, LVMi: Left ventricular mass indexed, LAVi: Maximal Left atrial volume indexed, LACI: left atrial-left ventricular coupling index. Continuous data are expressed as mean±SD or median [interquartile] values.

Table 3 details the 4D-Flow MRI-derived parameters. Peak velocities (E_TL_, A_TL_, and E/A_TL_) at the MV-TL did not differ between groups. However, V_P_ was markedly faster in HCM patients compared to controls (100.4 vs. 60.2 cm/s, p<0.01), a difference that remained significant after normalization by PEFR (p<0.01). While absolute PEFR were similar, the volume-normalized peak early filling rate (PEFR/FV) was significantly reduced in HCM (3.5 vs. 4.2 s⁻¹, p<0.01). Importantly, positive E IVPD and the absolute magnitude |E-ER| IVPD were significantly higher in the HCM group, whereas reverse ER IVPD was higher without reaching statistical significance (p=0.091). These differences were amplified when positive, absolute and reverse IVPDs were normalized by PEFR (p<0.001, p<0.001, and p<0.01, respectively). Decomposition of the pressure gradients revealed that the group differences were driven primarily by the inertial components of the IVPD, whereas the convective terms remained similar.

**Table 3:**
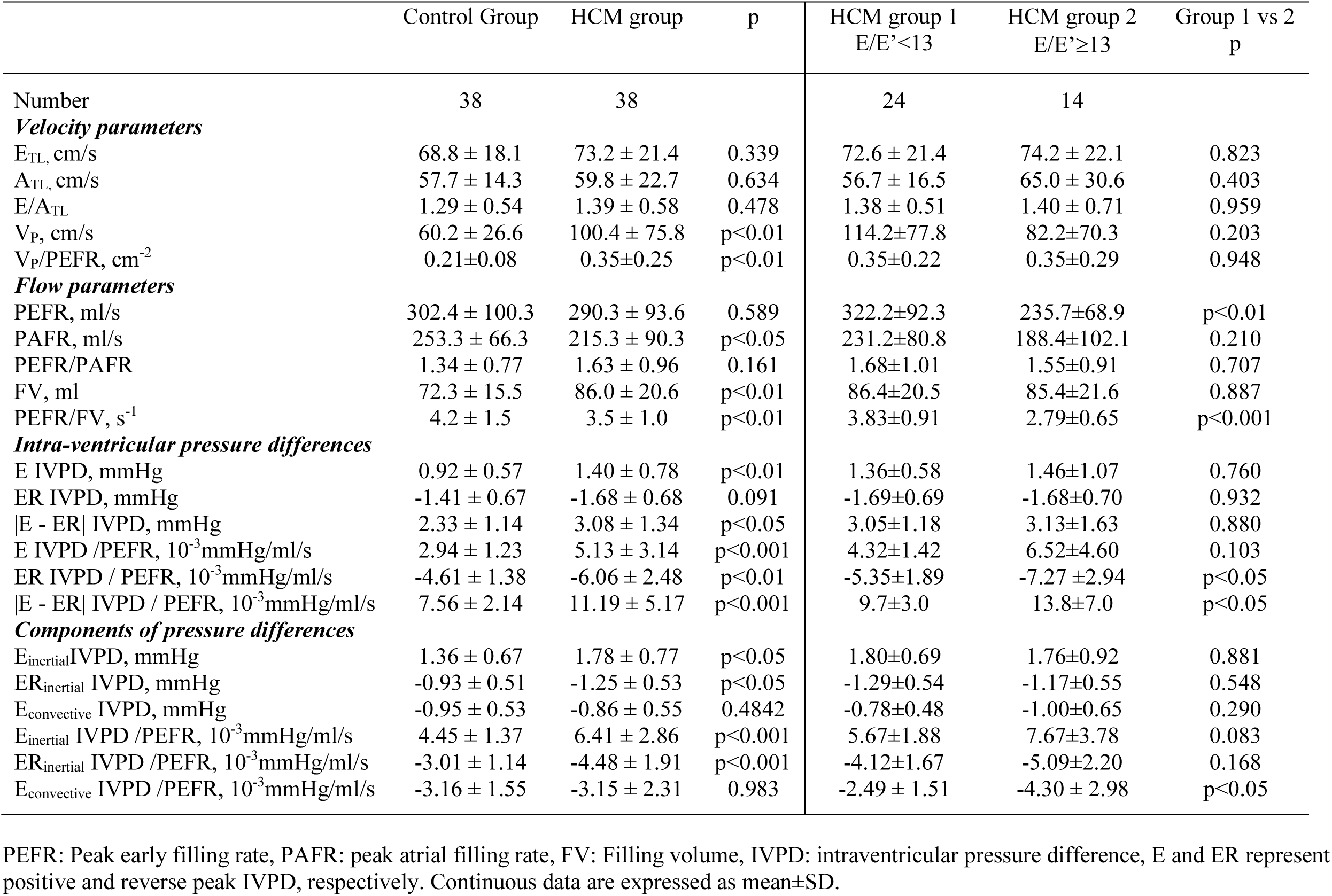
MRI 4D-Flow derived parameters including Vp and IVDPs in control group, HCM group and subgroups of HCM based on an TTE E/E’ cutoff of 13.

When stratifying the HCM cohort by TTE in subgroup 1 with E/E’<13 (n=24, 63%) and subgroup 2 with E/E’≥13 (n=14, 37%) Table 1 shows that weight, body mass index and BSA were higher in HCM subgroup 1 than in subgroup 2 (p<0.01, p<0.05, and p<0.01, respectively). BNP was higher in HCM subgroup 2 than in subgroup 1 (p<0.01). Table 2 only shows differences in TTE measurements between the HCM subgroups for E_TTE_ (p<0.05). Data from the MRI measurements showed significantly higher LVMi values in cases of E/E’≥13 (p<0.05), along with a similar upward trend for LAVi (p=0.50) and LACI (p=0.068), with no significant extent of LGE.

Table 3 shows no difference in 4D-Flow velocity measurements between the two HCM subgroups. However, 4D-Flow data show a decrease in PEFR and the PEFR/FV ratio in cases of diastolic dysfunction (p<0.01 and p<0.001, respectively). Regarding IVPDs, only the magnitude of ER IVPD and |E - ER| IVPD were greater in HCM patients with E/E’ ≥13, and only when both were normalized by PEFR (p<0.05). Figure 2 illustrates representative IVPD color maps and time curves across the clinical spectrum. As shown in Supplementary Tables 1 and 2 and illustrated in Figure 3, although most IVPD magnitudes increased progressively alongside worsening LAVi or LACI, none of the IVPD indices could reliably distinguish between HCM subgroups based solely on these structural severity thresholds or BNP levels.

**Figure 2:**
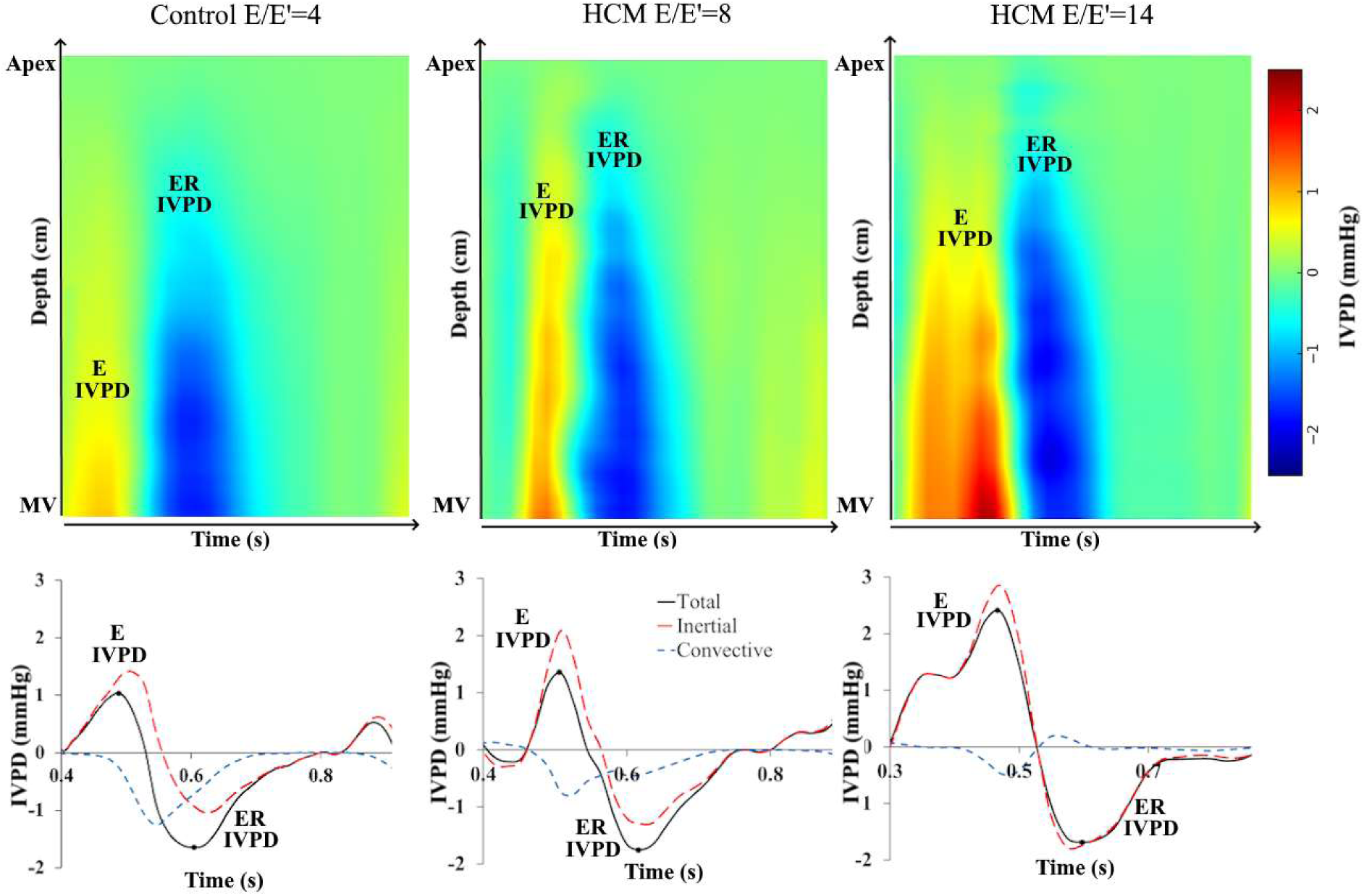
Representative examples of intraventricular pressure differences (IVPDs) in a healthy control and two HCM patients. The HCM patients represent subgroups with an echocardiographic E/E’ ratio < 13 and ≥ 13. **(Top)** Spatiotemporal IVPD color maps extending from the mitral valve tip leaflets (MV-TL, bottom) to the left ventricular apex (top) across the cardiac cycle. **(Bottom)** Corresponding temporal curves detailing the total IVPD along with its inertial and convective components. E-IVPD and ER-IVPD denote the peak positive and peak reversal pressure differences, respectively.

**Figure 3:**
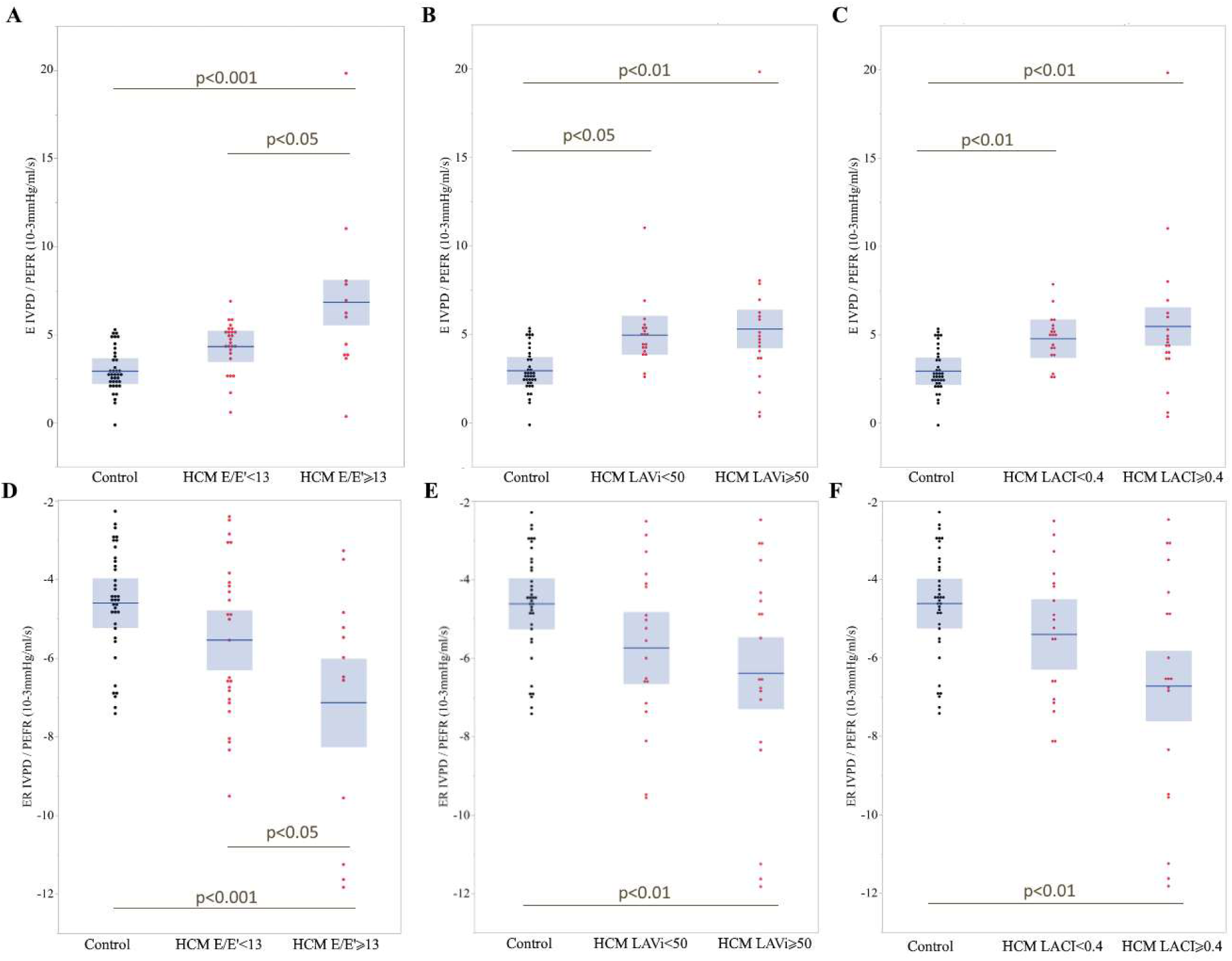
Boxplots representing positive (E IVDP) and reversal (ER IVPD) pressure difference values normalized by PEFR for controls and patients with HCM. HCM patients were stratified into two subgroups based on echocardiographic E/E’ (panels **A** and **D**), indexed left atrial volume (LAVi) (panels **B** and **E**), and left atrioventricular coupling index (LACI) (panels **C** and **F**).

Table 4 shows the univariate correlations with the main determinants of positive and reversal IVPDs before and after PEFR normalization. The strongest associations for such IVPDs were found with 4D-Flow measurements. E IVPD was found related to E_TL_ (r=0.45), PEFR (r=0.37) and V_P_ (r=0.65), all with p<0.001. The same variables showed significant associations for reversal ER IVPD (r=-0.81, r=-0.58 and r=-0.40, respectively; all p<0.001). The E/A_TL_ and PEFR/PAFR ratios were also associated with reversal IVPD ER (r=-0.53 and r=-0.52, respectively), with p<0.001. After normalization by PEFR, the positive E IVPD was associated with E/E’_TTE_ (r=0.35, p<0.01), LVM/LVEDV (r=0.30, p<0.01) and with V_P_ (r=0.47, p<0.001), whereas ER IVPD correlated with E/E’_TTE_ (r=-0.42, p<0.001) and E_TL_ (r=-0.44, p<0.001).

**Table 4.** Correlations between intra-ventricular pressure differences (IVPDs) and TTE, MRI and 4D-Flow measurements for the entire cohort (n=76).

|  | E IVPD<br>r value | p | ER IVPD<br>r value | p | E IVPD<br>/PEFR<br>r value | p | ER IVPD /PEFR<br>r value | p |
| --- | --- | --- | --- | --- | --- | --- | --- | --- |
| Age | -0.29 | 0.01 | 0.36 | <0.01 | 0.04 | 0.76 | -0.09 | 0.45 |
| **TTE**<br>**measurements** |  |  |  |  |  |  |  |  |
| E<sub>TTE</sub> | 0.24 | 0.04 | -0.32 | <0.01 | 0.25 | 0.03 | -0.40 | <0.001 |
| E/A<sub>TTE</sub> | 0.23 | 0.05 | -0.35 | <0.01 | 0.05 | 0.70 | -0.08 | 0.53 |
| E<sub>TTE</sub>/E' | 0.06 | 0.62 | 0.01 | 0.93 | 0.35 | <0.01 | -0.42 | <0.001 |
| **MRI**<br>**measurements** |  |  |  |  |  |  |  |  |
| LVEF | 0.28 | 0.01 | -0.22 | 0.05 | 0.28 | 0.02 | -0.29 | 0.01 |
| LVEDVi | 0.05 | 0.69 | -0.08 | 0.50 | -0.14 | 0.24 | 0.23 | 0.04 |
| LVESVi | -0.14 | 0.22 | 0.09 | 0.44 | -0.21 | 0.08 | 0.24 | 0.04 |
| LVMi | 0.25 | 0.03 | -0.19 | 0.10 | 0.22 | 0.05 | -0.22 | 0.05 |
| LVM/LVEDV | 0.22 | 0.06 | -0.18 | 0.11 | 0.30 | <0.01 | -0.38 | <0.001 |
| LAVi, ml/m<sup>2</sup> | -0.07 | 0.58 | 0.02 | 0.84 | 0.07 | 0.56 | -0.14 | 0.24 |
| LAVmin ml | -0.05 | 0.70 | 0.05 | 0.64 | 0.06 | 0.59 | -0.06 | 0.60 |
| LACI | -0.03 | 0.82 | 0.04 | 0.74 | 0.16 | 0.17 | -0.22 | 0.06 |
| **4D-Flow**<br>**measurements** |  |  |  |  |  |  |  |  |
| E<sub>TL</sub> | 0.45 | <0.001 | -0.81 | <0.001 | 0.15 | 0.21 | -0.44 | <0.001 |
| E/A<sub>TL</sub> | 0.24 | 0.04 | -0.53 | <0.001 | -0.03 | 0.80 | -0.11 | 0.38 |
| PEFR | 0.37 | <0.001 | -0.58 | <0.001 | - | - | - | - |
| PEFR/PAFR | 0.31 | <0.01 | -0.52 | <0.001 | - | - | - | - |
| PEFR/FV | 0.33 | <0.01 | -0.41 | <0.001 | - | - | - | - |
| V<sub>p</sub> | 0.65 | <0.001 | -0.40 | <0.001 | 0.47 | <0.001 | -0.21 | 0.07 |

TTE: Transthoracic echocardiogram, LVEF: Left ventricular ejection fraction, LVEDVi: Left ventricular end-diastolic volume indexed, LVESVi: Left ventricular end-systolic volume indexed, LVMi: Left ventricular mass indexed, LAVi: Left atrial volume indexed, LACI: left atrial-left ventricular coupling index. Continuous data are expressed as mean±SD or median [interquartile] values.

Figure 4 presents the multivariable models identifying the independent determinants of IVPDs. Model 1 (r = 0.50, p<0.001) demonstrates that E IVPD is independently driven by PEFR (p<0.001) and E/E’_TTE_ (p<0.01), regardless of age or LAVi. Model 2 (r = 0.65, p < 0.001) confirms that ER IVPD is similarly determined by PEFR (p<0.001) and E/E’_TTE_ (p<0.01), independent of age and LAVi. Figure 5 illustrates the high correlation found between ER IVPD and PEFR (r=-0.58; p<0.001).

**Figure 4.**
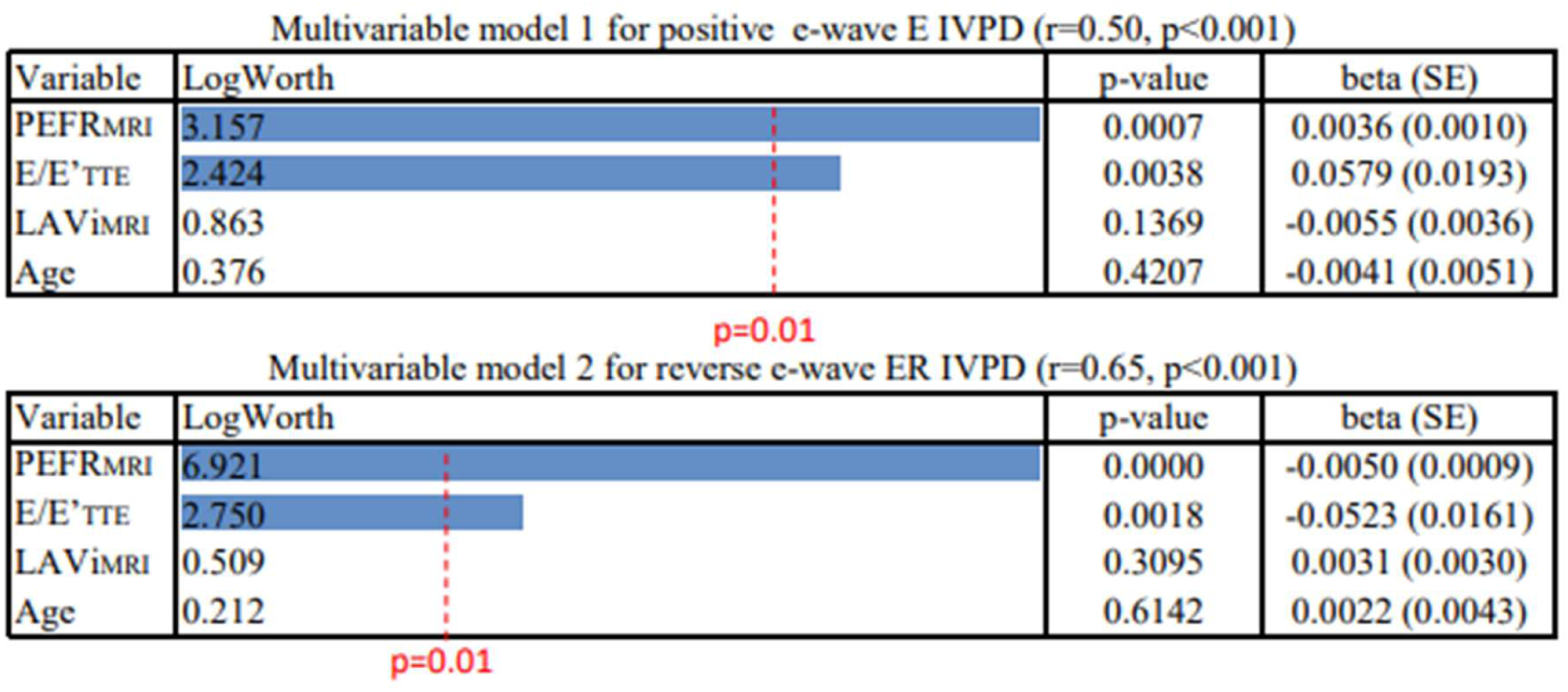
Multivariable regression models identifying the independent determinants of positive (E IVPD) and reversal (ER IVPD) intraventricular pressure differences. The models are adjusted for age, including peak early filling rate (PEFR), echocardiographic E/E’ ratio, and indexed left atrial volume (LAVi) as covariates.

**Figure 5.**
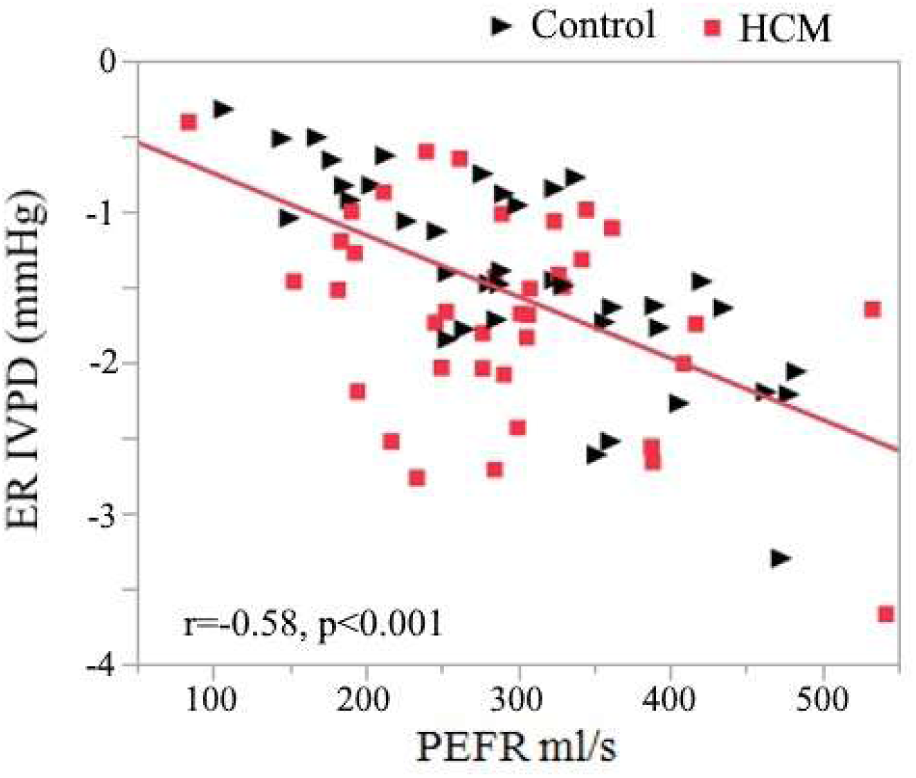
Correlation between the reversal intraventricular pressure difference (ER IVPD) and peak early filling rate (PEFR). Scatter plot demonstrating a strong inverse relationship (r = -0.58, p < 0.001) between 4D-Flow MRI-derived PEFR and ER IVPD across the entire study cohort. Healthy controls are represented by black triangles and HCM patients by red squares.

Finally, interobserver agreement was excellent for all primary 4D-Flow metrics, yielding ICCs of 0.997 [0.993-0.999] for E IVPD, 0.997 [0.993-0.999] for ER IVPD, and 0.999 [0.998-1.000] for Vp, with very low coefficients of variation (2.8%, 2.2%, and 1.8%, respectively).

## DISCUSSION

### Main Findings

In this study, we demonstrated the clinical feasibility and added value of using 4D-Flow MRI to non-invasively assess LV diastolic IVPD and V_P_ in patients with HCM. The principal findings of our work are: 1) 4D-Flow derived IVPD magnitudes (specifically the early positive E IVPD and the reverse ER IVPD) and V_P_ are significantly elevated in HCM patients compared to age- and sex-matched healthy controls; 2) the discriminatory power of these markers is noticeably increased when IVPDs are normalized by the PEFR; and 3) the magnitude of the reversal gradient ER IVPD is independently determined by both PEFR and the echocardiographic E/E’ ratio (a surrogate for LV filling pressure), regardless of patient age or LA size.

### Physiological Interpretation of IVPDs in HCM

The assessment of IVPDs provides unique insights into the delicate interplay between LA driving pressure and LV suction forces during early diastole. Physiologically, rapid active LV relaxation and elastic recoil generate a potent suction force, driving a base-to-apex pressure gradient. Because active relaxation is inherently impaired in HCM, one might theoretically expect this early suction gradient to be blunted[6,9,21,22]. Paradoxically, we found that both the positive E IVPD and the reverse ER IVPD were significantly higher in the HCM cohort. This increase is likely driven by elevated LA pressure forcefully compensating for the stiff, non-compliant LV. Our multivariable analysis firmly supports this finding, demonstrating an independent association between IVPD with both E/E’_TTE_ and PEFR. This aligns with physiological concepts suggesting that, in states of diastolic dysfunction, the LA to the mid LV (basilar) IVPD can be maintained or even increased by high LA pressure, even if true apical suction is severely compromised or lost[23]. Furthermore, this is consistent with prior experimental data (in humans and animals with normal LV function) showing that invasive IVPDs, as measured by catheterization, were found to be strongly correlated with LA pressure and sonomicrometry-based measurements of PEFR and of LV filling volume[23]. Interestingly, prior echocardiographic studies evaluating early filling intraventricular pressure gradients have frequently reported reduced gradients in patients with heart failure [21], severe HCM[21,24], or dilated cardiomyopathy[20]. In contrast, our 3D methodology revealed significantly increased gradients in HCM, particularly in cases with severe DD (E/E’≥13). By mathematically decomposing the gradients, we discovered that this overall IVPD increase is primarily driven by the inertial component (temporal acceleration of flow/velocity), mirroring the hemodynamics observed in restrictive mitral filling patterns (E/A > 2). Our findings are also consistent with a recent 2D planar phase-contrast MRI study in patients with HFpEF, which reported a marked increase in the amplitude of the reverse ER IVPD compared to controls, though that study did not establish a link with standard echocardiographic DD criteria[22].

### The Value of Flow Normalization (IVPD/PEFR)

A novel and highly discriminant aspect of our methodology is the normalization of IVPDs by PEFR, which is the primary determinant of the IVPDs. When normalized to flow, the absolute differences in IVPD between the HCM cohort and healthy controls, as well as between HCM severity subgroups (E/E’ < 13 vs. ≥ 13), became highly significant. This metric captures the core pathophysiology of HCM: for any given unit of blood flow entering the ventricle, the required pressure difference generated to move that blood is substantially higher (Figure 6). Therefore, flow-normalized IVPD is a highly sensitive, flow-independent marker of underlying chamber stiffness and impaired diastolic compliance, both of which are significantly impaired in HCM[25]. This accounts for the increased slope of LV pressure-volume curves during diastole as previously reported in cases of heart failure with preserved ejection fraction (HFpEF)[26].

**Figure 6:**
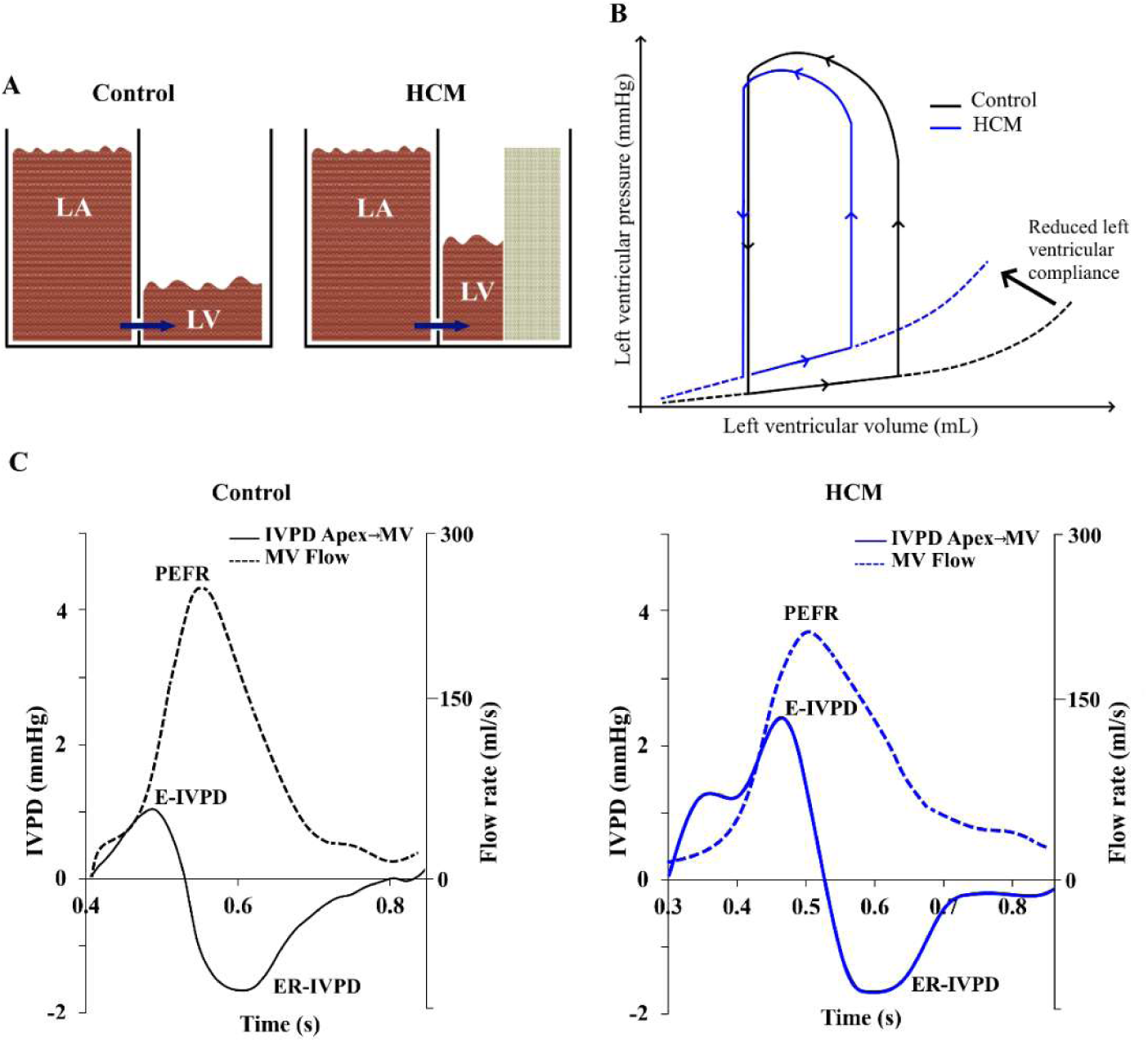
Proposed hemodynamic framework linking classical left ventricular diastolic compliance to 4D-Flow MRI-derived pressure gradients. **(A) Hydrodynamic analogy of diastolic filling.** A communicating-cardiac chamber model illustrating left atrial (LA) to left ventricular (LV) volume transfer. In hypertrophic cardiomyopathy (HCM, right), the thickened LV wall (gray block) restricts chamber capacitance. Consequently, driving a similar fluid volume into the restricted LV requires a disproportionately higher hydrostatic pressure gradient from the LA compared to a normal, compliant LV cavity (Control, left). **(B) Classical Pressure-Volume (P-V) relationship.** Schematic P-V loops demonstrating the physiological basis of diastolic dysfunction. The HCM heart (blue) exhibits a leftward shift and a steepened diastolic pressure-volume relationship compared to the healthy control (black). This steeper slope indicates reduced diastolic compliance, wherein a small physiological change in volume induces an exaggerated increase in pressure. **(C) *In vivo* translation using 4D-Flow MRI.** Representative temporal profiles of mitral valve inflow rate (dashed lines, right axis) and the resulting intraventricular pressure differences (IVPD, solid lines, left axis) during early diastole. While the HCM patient (right) exhibits a similar or slightly reduced peak early filling rate (PEFR) compared to the control (left), generating and decelerating this flow requires markedly amplified pressure gradients (manifested by a higher positive E IVPD and a deeper reversal ER IVPD). This visually underscores how normalizing IVPD by PEFR (IVPD/PEFR ratio) effectively captures the impaired compliance and chamber stiffness depicted in Panels A and B.

Previous ultrasound studies have also consistently shown that early filling IVPDs increase following an increase in underlying flow (e.g., during exercise[21], post-revascularization[8], or post-myectomy in HCM[23]). This reinforces our premise that pressure gradients must be interpreted in the context of their underlying flow rate, a concept well-established in the assessment of aortic stenosis[27,28] and coarctation[29].

When we consider the LAVi and LACI (two other indirect criteria reflecting an increase in LV diastolic pressures[7,17]) we find more modest, or even non-significant, associations with increased IVPDs. These indirect criteria, which are based primarily on an increase in LAV, are not solely related to elevated LV pressures, as other independent factors influence this atrial dilation (e.g. presence of atrial fibrillation and/or mitral regurgitation). Further studies are needed to compare the diagnostic and prognostic relevance of IVPDs with these indices, which can be easily measured using MRI.

### Propagation Velocity (V_P_) Re-evaluated by 4D-Flow

Our data demonstrate that V_P_ is significantly higher in patients with HCM than in controls. However, the literature on V_P_ in HCM patients using CDMM echocardiography is conflicting, with some studies reporting a decrease[23] and others an increase[25]. The determinants of V_P_ are linked to peak transmural velocity, filling volume, and complex ventricular geometry[12,25]. In HCM patients, the severe reduction in cavity size and asymmetric septal thickening can unpredictably alter the convective acceleration of incoming blood. The 1D nature of CDMM makes it exceptionally vulnerable to scan-line misalignment and spatial under-sampling. If the inflow jet is skewed (a near-certainty in the distorted geometry of HCM) CDMM will miss the true peak velocity vector[9]. 4D-Flow MRI circumvents these geometric limitations by capturing the entire 3D velocity field, allowing for a more precise tracking of the true flow pattern. Physiologically, the elevated V_P_ observed in our 4D-Flow data likely reflects the forceful, high-pressure injection of blood from a hypertensive LA into a restrictive, non-compliant LV cavity. However, in our cohort, flow-normalized IVPDs outperformed V_P_ in distinguishing HCM patients from controls and in discriminating severe DD (E/E’ ≥ 13).

### Advantages of 4D-Flow over 2D-Flow MRI or CDMM Echocardiography Methods

While 2D phase-contrast MRI and CDMM echocardiography have been validated for IVPD estimation, they remain constrained by planar or linear assumptions. Computational fluid dynamics (CFD) models have proven that incorporating multidimensional velocity information significantly improves the accuracy of IVPD calculations, particularly in mid-to-late diastole when complex multidimensional vortex formations occur[10]. By using whole-heart 4D-Flow MRI, our approach better accounts for the complex 3D pathways inherent to LV filling, preventing the underestimation of flow velocities caused by improper 1D or 2D alignment[22].

### Limitations

Our study has several limitations. First, it is a single-center study with a moderate sample size, though the groups were carefully matched. Second, we did not perform simultaneous invasive cardiac catheterization to serve as a gold-standard comparison for our 4D-Flow derived IVPDs. However, the theoretical basis of calculating non-invasive gradients via the Navier-Stokes and Euler equations is well established and has been validated in previous models with MRI[6,10,27,28,29]. Third, the temporal resolution of our 4D-Flow sequence is lower than that of continuous-wave or CDMM echocardiography. While temporal resolution is a critical parameter for calculating the partial derivative terms in the Euler equation, previous studies have demonstrated that down-sampling MRI temporal resolution (e.g., to ∼14 ms) has a minimal impact on the peak values of major IVPD waves[9,28].

## Conclusion

In conclusion, 4D-Flow MRI can quantify diastolic IVPDs and propagation velocity V_P_ in patients with hypertrophic cardiomyopathy. Elevated, flow-normalized IVPDs strongly correlate with traditional markers of elevated left ventricular filling pressures, offering a deeper, comprehensive 3D hemodynamic characterization of diastolic dysfunction and chamber stiffness.

## Data Availability

The data mentioned in the manuscript and in our database may eventually be shared, subject to approval by the head of the REMY registry, Prof. A. Hage, a co-author of our study, who must be consulted in such cases.

## Acknowledgments

We would like to thank the French Society of Cardiology, the sponsor and host of the database of the REMY registry.

